# Investigating the Causal Relationship Between Sleep-Related Traits and Self-Reported Diabetes: A Mendelian Randomization Study

**DOI:** 10.1101/2024.09.09.24313314

**Authors:** Nismabi A Nisamudheen, Dinesh Velayutham, Puthen Veettil Jithesh

**Author notes:** Corresponding Author: Puthen Veettil Jithesh, PhD FRSB, College of Health & Life Sciences, Hamad Bin Khalifa University, Qatar Foundation, C-147, Penrose House, PO Box 34110, Education City, Doha, Qatar, E.

## Abstract

**Objective:** Self-reported data can be a valuable resource for understanding health outcomes, behaviors, disease prevalence, and risk factors, yet underutilized in epidemiological research. While observational studies have linked sleep traits with diabetes, evidence using self-reported diabetes data for causal connection is lacking.

**Methods:** We performed a two-sample Mendelian randomization (MR) analysis using Inverse Variance Weighting (IVW), IVW with multiplicative random effects (IVW-MRE), Maximum Likelihood (ML), MR-Egger regression, and Weighted Median models, with genetic variants linked to five sleep traits (sleep duration, insomnia, snoring, daytime dozing, and chronotype) and self-reported diabetes from the UK Biobank dataset. The study utilized MR-Egger and MR-PRESSO regression to evaluate pleiotropy and outliers, IVW Q statistics to detect heterogeneity, the MR-Steiger test to assess directionality, and leave-one-out sensitivity analysis to ensure the reliability.

**Results:** ML provided positive causal associations between genetically predicted insomnia (p = 0.002, OR = 1.021, 95% CI: 1.008–1.035) and daytime dozing (p = 0.014, OR = 1.029, 95% CI: 1.006–1.052) with diabetes, while IVW and IVW-MRE analysis showed a trend towards significance. Snoring showed mixed evidence, while genetically predicted sleep duration was marginally associated with diabetes (p = 0.053, OR = 0.992, 95% CI: 0.984–1.000) with the weighted median method, indicating a potential small protective effect. No causal association was found between chronotype and diabetes.

**Conclusion:** This exploratory MR study provides evidence for the effect of insomnia, daytime dozing, sleep duration and snoring on diabetes risk. These findings underscore the importance of considering self-reported health outcomes in epidemiological research.

**Article Highlights:** *Why did we undertake this study?:* We undertook this study to explore the causal relationships between sleep-related traits and diabetes using self-reported data, as previous prospective, retrospective or other observational studies have shown associations but lacked causal evidence using self-reported data.

*What is the specific question(s) we wanted to answer?:* We aimed to answer whether sleep-related traits, such as sleep duration, insomnia, snoring, daytime dozing, and chronotype, have a causal impact on diabetes which is self-reported.

*What did we find?:* Our two-sample Mendelian Randomization (MR) analysis found that genetically predicted insomnia and daytime dozing have a positive causal association and sleep duration was marginally associated with diabetes.

*What are the implications of our findings?:* These findings suggest that certain sleep traits may contribute to diabetes risk, highlighting the importance of considering sleep in diabetes prevention and treatment strategies. The results also emphasize the value of using self-reported health outcomes in epidemiological research and clinical interventions.

## Background

Diabetes is a chronic metabolic disease marked by elevated blood sugar levels. In 2019, diabetes was estimated to affect 9.3% of the global population, impacting around 463 million people. Projections indicate that by 2030, this prevalence could increase to 10.2% (578 million people) and further rise to 10.9% (700 million people) by 2045[1]. The prevalence of obesity has been rising as well, with obesity and diabetes becoming major public health concerns [2], emphasizing the importance of understanding the characteristics and risk factors associated with these diseases [3].

Sleep plays a crucial role in metabolic health and the development of diabetes. Recent research has focused on understanding the genetic underpinnings of sleep behaviors and their impact on diabetes risk. A recent study using genome-wide association analyses to identify new loci associated with sleep duration and chronotype offered insights into the genetic regulation of sleep patterns [4]. Meanwhile, genetic studies of accelerometer-based sleep measures were employed to uncover the molecular mechanisms underlying sleep behaviors and their potential causal links with chronic diseases like diabetes and obesity [5].

Mendelian randomization (MR) is an epidemiological technique that evaluates the causal links between modifiable exposures and outcomes, such disease status, by using genetic variants as instrumental variables. MR utilizes genetic variants as instrumental variables to evaluate the causal effects of exposures, such as sleep behaviors, on health outcomes like diabetes. The fundamental principle of MR is based on genetic variants that are randomly allocated at conception, resembling a randomized controlled trial setting, which helps mitigate confounding and reverse causation biases often present in observational studies. The key assumption of MR is that the genetic instruments are linked to the exposure of interest and affect the outcome exclusively through that exposure, thereby enabling a valid causal inference.

Recent studies have employed MR to identify causal risk factors for diabetes as well as to explore the genetic basis of sleep traits [6, 7]. By utilizing genetic data from genome-wide association studies (GWAS), MR investigations can elucidate the causal relationships between sleep patterns, genetic variants, and the risk of diabetes. Findings from MR analyses can guide public health strategies, personalized interventions, and further research into the mechanisms linking sleep traits to diabetes outcomes.

Self-reported data, often collected through surveys or questionnaires, provide valuable insights into an individual’s health conditions, behaviors, and needs that might not be captured through clinical or administrative data alone. Studies have emphasized the importance of self-reported health outcomes in diabetes management and research [8][9]. A recent study on factors related to oral health literacy among patients with type 2 diabetes, stressed the importance of enhanced health literacy in diabetes management [10] and another research assessed the validity and reliability of self-reported diabetes in the Atherosclerosis Risk in Communities Study, underscoring the significance of accurate self-reports for epidemiological research [11].

Despite the growing body of research on the associations between sleep traits and diabetes, there remains a research gap in comprehensively investigating the causal relationship between sleep-related traits as exposures and diabetes as outcome using self-reported data. Therefore, we investigated the causal link between various sleep-related traits (sleep duration, insomnia, chronotype, snoring, and daytime dozing) as exposure and diabetes as the outcome. We specifically used self-reported diabetes as the outcome to investigate the reliability of self-reported data in replicating the results obtained using measured clinical parameters that define diabetes.

## Methods

### Study Design

The study workflow is shown in Figure 1. First, we extracted the instrumental variables (IVs) for the exposure from the GWAS database. In this study, we used various sleep-related traits as exposure, including sleep duration, insomnia, chronotype, snoring, and daytime dozing. Second, we extracted SNPs from the GWAS study of self–reported diabetes that matched the IVs. Thirdly, we harmonized the GWAS data for the exposures and outcomes to ensure the influence of IVs through exposure on outcomes. Then we calculated F statistic and R^2^ (proportion of variance of exposure explained by the IVs) to evaluate the strength of the chosen SNPs. Fourth, we conducted MR analysis to examine the causal relationship between sleep traits and self-reported diabetes. We also performed pleiotropy, heterogeneity, and outlier tests to identify whether the causality is reliable and robust. Next, we performed the MR Steiger directionality test to identify the correct direction of the causal effect between exposures and outcomes. Outliers identified have been removed to improve the validity of causal inference in our MR studies. Again, we repeated our MR analysis using the residual IVs to establish the causal association. Finally, we conducted Single SNP Analysis and leave-one-out analysis to obtain a more robust and reliable causal relationship.

**Figure 1:**
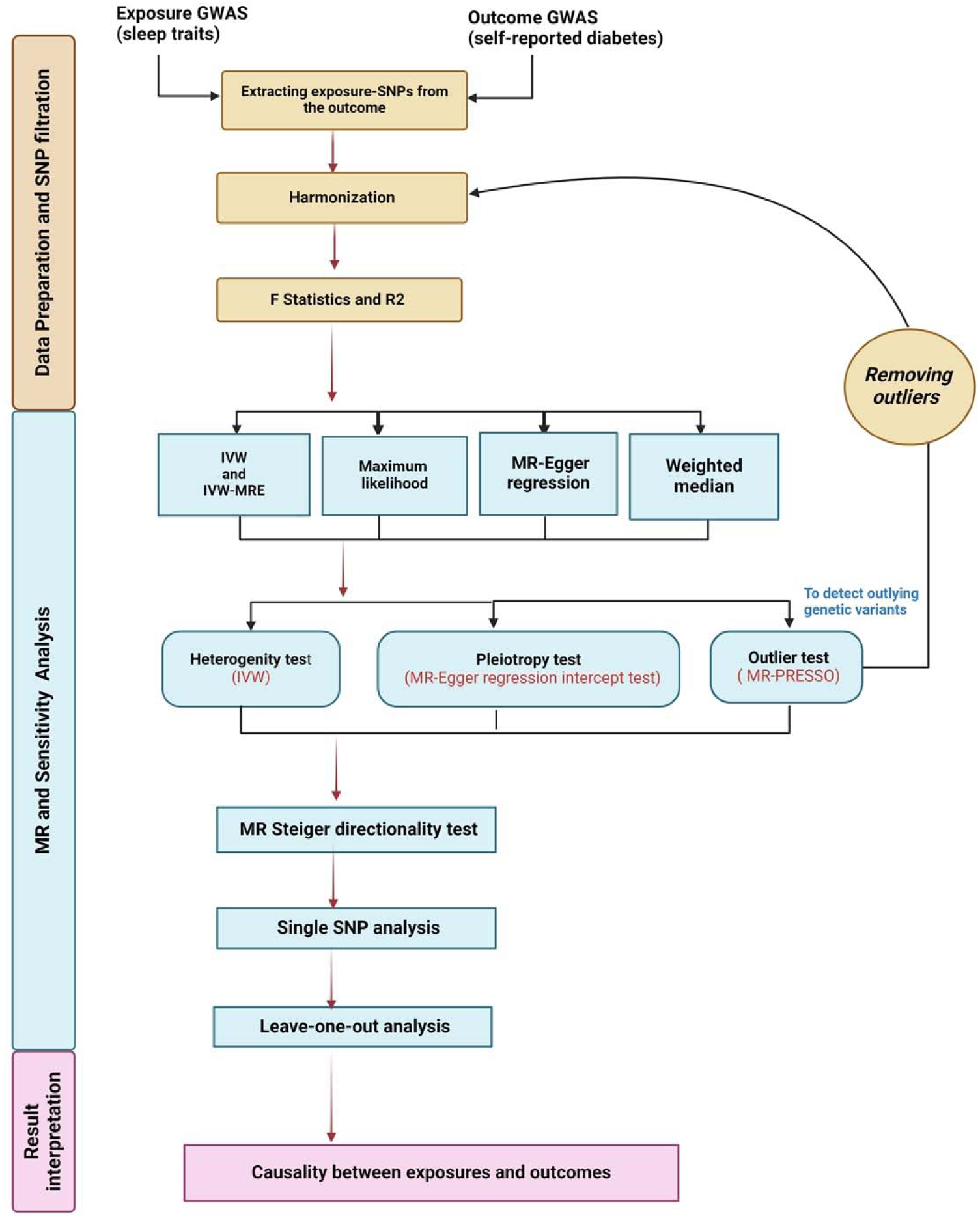
Flow chart for the Mendelian Randomization (MR) study

### Data Sources

We utilized publicly available GWAS datasets to obtain instrumental variables. Our exposure and outcome data came from the IEU Open GWAS project database (https://gwas.mrcieu.ac.uk/), a large research cohort with over 500,000 participants from throughout the UK. Two self– reported diabetes outcome datasets were extracted from Non-cancer illness code: Self-report, UK Biobank data. Complete information on the selected GWAS datasets is shown in Supplementary File 1 Table 1.

### Selection of Instrumental variable

Genome-wide significance (P < 5 x 10□^8^) was used as the threshold to select IVs for the exposure. However, if there were less than 10 SNPs, we used relaxed p-value threshold (P < 5 x 10□^5^) to confirm prior findings and potentially uncover new insights specific to self-reported diabetes. This threshold was chosen to enhance the ability to replicate these associations in independent samples with self-reported diabetes, based on prior strong evidence from non-self-reported diabetes studies. The clumping distance threshold was set to 10,000 kb, while filtering those in linkage disequilibrium LD-r² = 0.001 [12]. We utilized the F statistic to assess the IVs’ strength, with weak IVs defined as F-statistics less than 10, and all weak IVs were deleted. Through matching IVs, SNPs in the exposure and outcome datasets were harmonized. Palindromic SNPs were excluded directly [12]. The SNPs remaining after these screening steps were considered the final instrumental variables representing the exposures and are detailed in Supplementary File 2.

Because the study was based on published data, ethical approval was not necessary for study under local legislation and institutional guidelines. Our work was reported according to the STROBE-MR reporting guidelines. [13].

### The Core MR Assumptions

Three fundamental assumptions based on the MR research design principles must be satisfied to obtain unbiased estimates; they are (i) SNPs chosen as IVs need to be strongly associated to exposure risk factors (sleep-related traits); (ii) SNPs are independent of confounding factors; (iii) genetic variation affects diabetes outcomes only through its effect on sleep-related traits (exposure).

### Statistical Analysis

Exposure variables were measured in units defined by the GWAS, while the outcome variable, diabetes, was considered as binary (presence or absence) and was self-reported. Genetic variants were utilized as IVs, and five methods were employed for conducting two-sample MR. Inverse Variance Weighting (IVW) method was used as the primary statistical method, to assess whether the genetic variants were valid instruments and to establishes statistical significance as p < 0.05 [14]. The other four procedures were employed for complimentary analyses. The random-effects IVW model was used to account for any heterogeneity. We also used the maximum likelihood model, which assumes no heterogeneity and horizontal pleiotropy. The MR-Egger regression strategy was also employed, that complements the IVW method by detecting and correcting for violations of IV assumptions, even when instruments are invalid [15, 16]. The Weighted Median method provides an unbiased estimate of causality even when up to 50% of the weight comes from invalid instrumental variables [16, 17].

We conducted multiple sensitivity analyses to evaluate the robustness of our findings against potential pleiotropy and heterogeneity in the causal estimates. These tests also assisted in identifying and addressing any influence from missing data or outliers on the accuracy of the MR estimates. First, we performed pleiotropy analysis using MR-Egger regression and evaluated the horizontal pleiotropy. Then we employed the MR-PRESSO outlier test to identify and remove the SNPs leading to horizontal pleiotropy. We obtained the empirical p value for the MR-PRESSO global test and when the corrected p value was not consistent with raw result, we removed all outliers from genetic instruments [18–20]. Secondly, to ascertain whether heterogeneity is present in the above-mentioned methods, we employed IVW method, with heterogeneity assessment using Cochran’s Q statistic [16, 21, 22]. To obtain MR estimates for each SNP individually, we did single SNP analysis using the Wald ratio approach that calculates the ratio of the SNP-outcome estimate to the SNP-exposure estimate [23]. To determine the orientation of the exposure’s effect on the outcome, we used MR Steiger directionality test [24]. Then leave-one-out sensitivity analysis utilizing the Wald technique was used to evaluate the influence of specific SNPs on the overall effect-size estimate and to identify influential variants [25].

All statistical analysis were performed using “TwoSampleMR” (version 0.5.7), “MRPRESSO” packages in R 4.3.1 (R Foundation for Statistical Computing, Vienna, Austria). Results are presented as odds ratios (OR), 95% confidence intervals (CI), and p-values with statistical significance was determined at p ≤ 0.05.

## Results

### Sleep-related Traits and Self-reported Diabetes

The summary association statistics extracted for the genome-wide significant SNPs associated with different sleep-related traits were matched and harmonized with that of diabetes to be for the same reference allele. Pleiotropy test performed using MR-Egger indicated negligible pleiotropy across all sleep traits (Supplementary file 1: Table 2). The Egger intercept values ranged from -0.0012 to 0.0003, with corresponding p-values exceeding 0.05, suggesting no evidence of directional pleiotropy. The results of the IVW heterogeneity tests showed significant heterogeneity for some sleep traits (Supplementary file 1: Table 2). Hence, SNPs in high LD with other SNPs and related to known confounders were excluded. All the outliers that were identified by MR-PRESSO Q test were removed from the instrumental set, which was confirmed in rerun of MR-PRESSO. No horizontal pleiotropy was detected based on MR-Egger intercepts; however, evident heterogeneity persisted despite these adjustments. (Supplementary File 1: Table 3).

In the reperformed MR analysis, chronotype (p = 0.655, OR = 1.001, 95% CI: 0.995–1.008) did not exhibit significant associations with diabetes risk, as indicated by beta values close to zero and p-values above 0.05. In contrast, snoring (beta = -0.048, p = 0.00013, OR = 1.007, 95% CI: 0.998–1.016) demonstrated a significant inverse association with diabetes risk, and all MR methods revealed consistent estimates suggesting a potential relationship. Notably, insomnia (p = 0.002, OR = 1.021, 95% CI: 1.008–1.035) and daytime dozing (p = 0.014, OR = 1.029, 95% CI: 1.006–1.052) showed a positive association with diabetes risk across the methods (Supplementary file 1 Table 4). However, sleep duration established marginal causal effect with weighted median method (p = 0.053, OR = 0.992, 95% CI: 0.984–1.000).

Overall, while insomnia and daytime dozing exhibited consistent positive causal associations, sleep duration did show a marginal impact on diabetes risk, while chronotype and snoring lacked clear associations. Figure 2 represents the forest plot for the overall causal association estimates between all sleep-related traits and self-reported diabetes. The MR Steiger test confirmed the causal direction of analysis (Supplementary file 1 Table 5). To investigate the effect of particular SNPs on the estimations, a single-SNP analysis and a leave-one-out analysis were used. The findings were consistent with those obtained when all accessible SNPs were included in the analysis.

**Figure 2.**
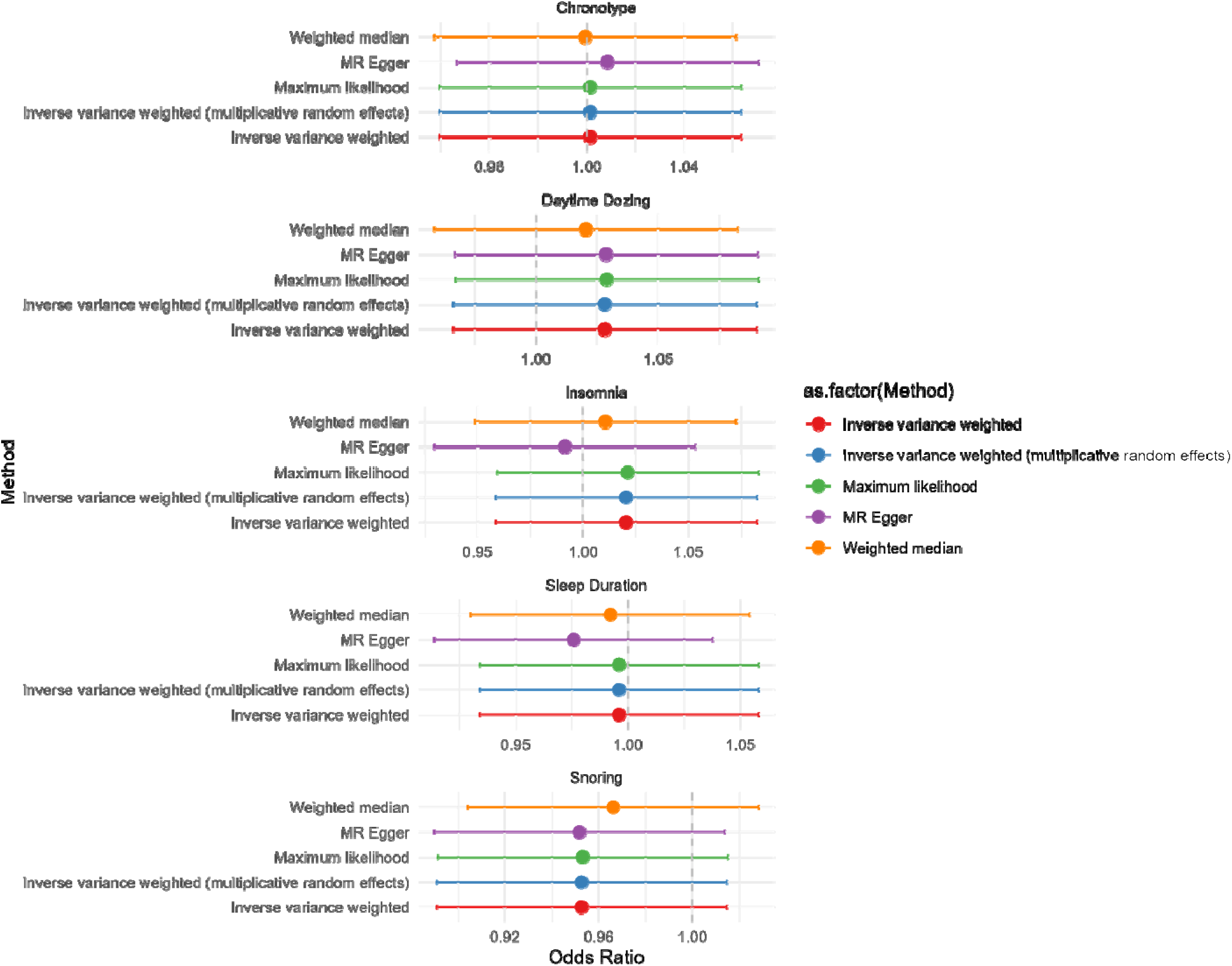
Association between genetically predicted sleep-related traits and self–reported diabetes

To ensure the reliability and clarity of our research findings, various graphical plots are employed to highlight potential biases, outliers, and the overall consistency of the data. Graphical plot representations for various analyses of insomnia and daytime dozing are depicted in Figures 3 & 4 and for others in Supplementary file Fig S1–Fig S3. The scatter plot visualizes the associations between SNPs and outcomes against SNP-exposure associations, providing a quick overview of the causal-effect estimates for each variant across different MR methods. The forest plot displays the effect sizes and confidence intervals of individual studies, helping to identify overall trends and assess heterogeneity. The leave-one-out sensitivity analysis systematically excludes each SNP, one at a time, and re-evaluates the causal effect estimate, ensuring that no single SNP disproportionately influences the overall result. Lastly, a funnel plot, a scatter diagram, is used to detect reporting bias in MR analysis by plotting the effect of exposure against study precision. If there is no bias, the plot should resemble a symmetrical funnel; any asymmetry may indicate the presence of bias.

**Figure 3:**
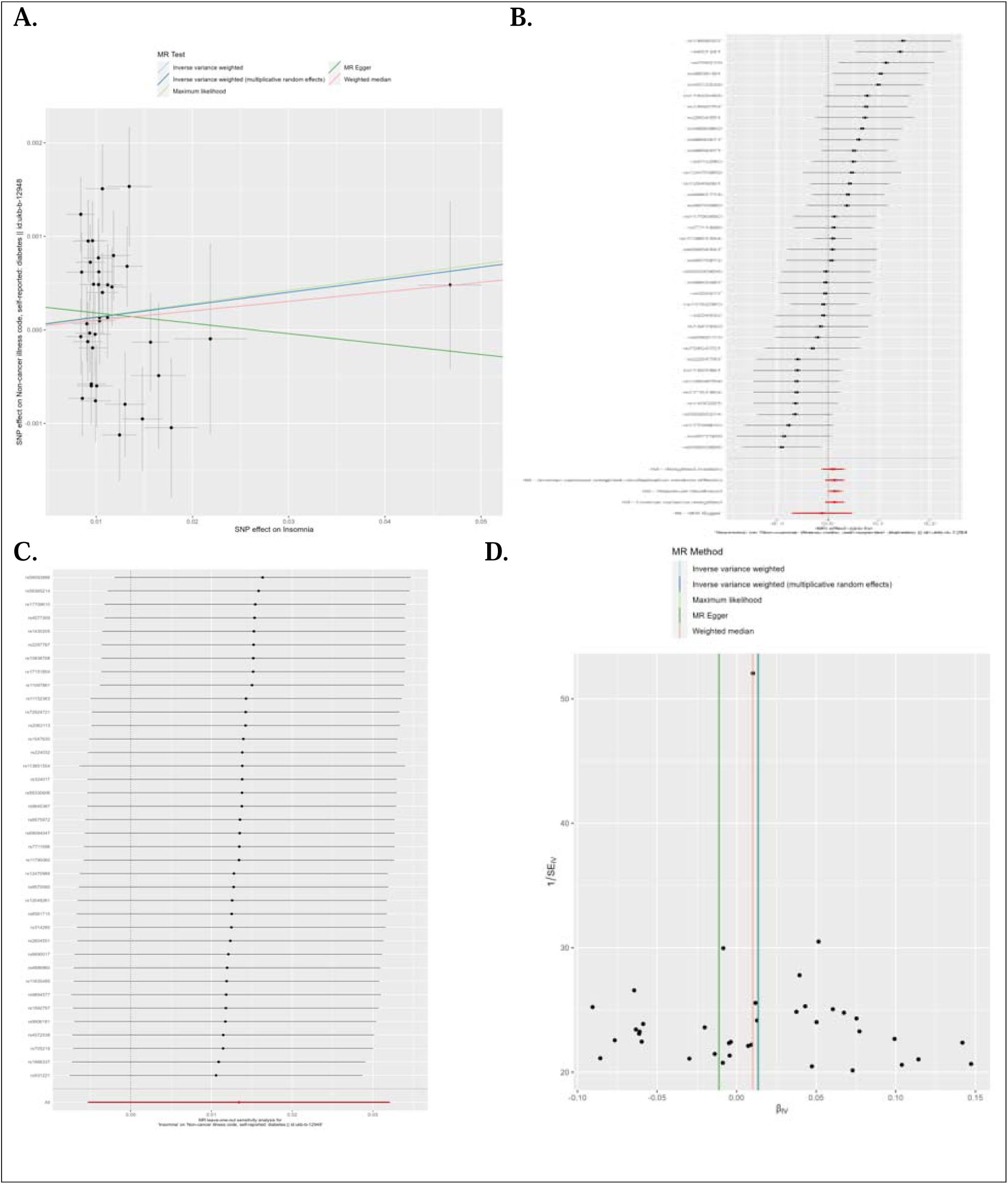
A. Comparison of results using different MR methods using scatter plot between Insomnia and Self –reported Diabetes B. Forest plot of single SNP MR C. Leave-one-out sensitivity analysis D. Funnel plot

**Figure 4:**
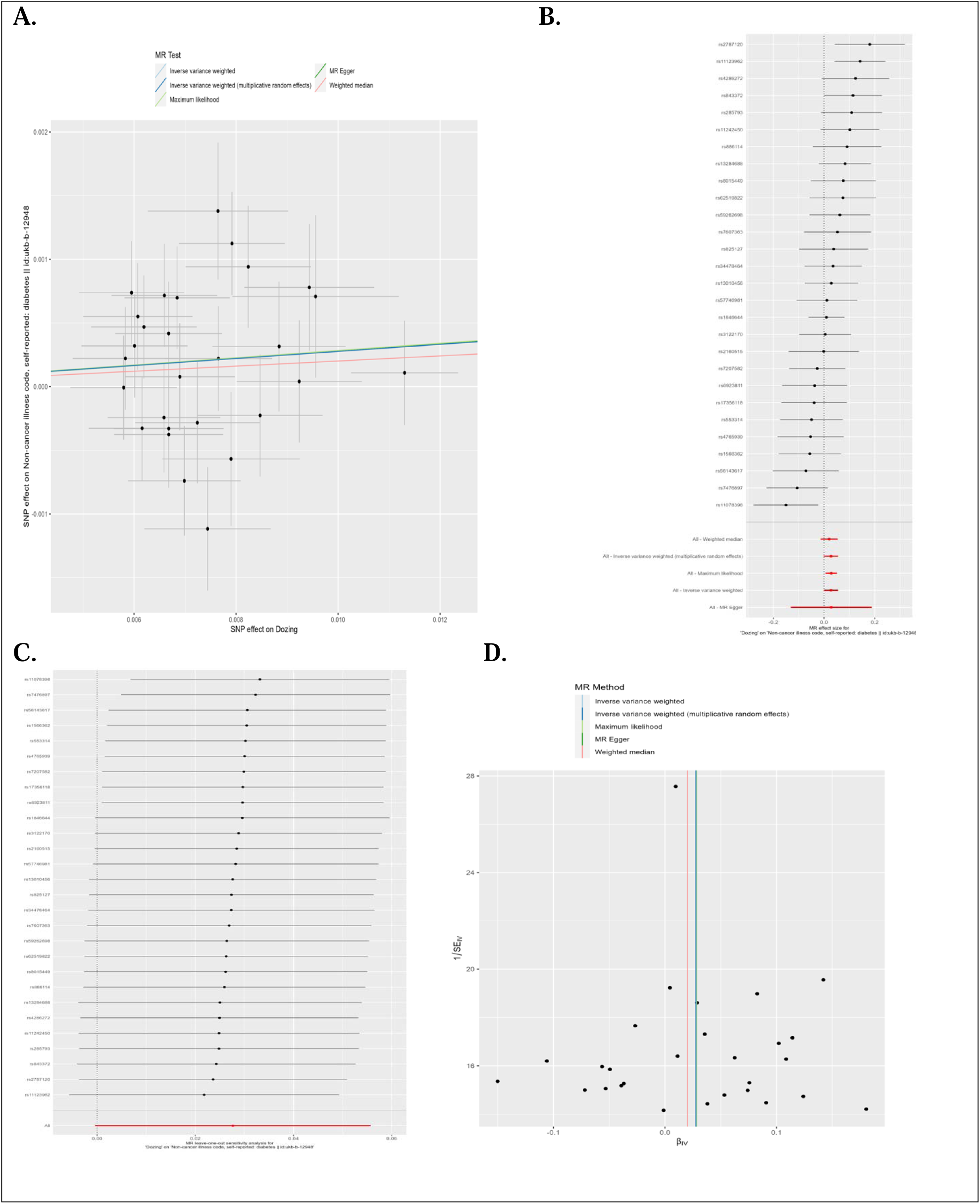
A. Comparison of results using different MR methods using scatter plot between Daytime dozing and Self –reported Diabetes B. Forest plot of single SNP MR C. Leave-one-out sensitivity analysis D. Funnel plot

## Discussion

In this study, we performed MR analysis to provide genetic evidence for causal relationship between sleep related traits and self-reported diabetes. Using self–reported diabetes dataset, we identified that insomnia and hypersomnia have potential positive associations, suggesting these problems may slightly increase the risk of diabetes. These results suggest that individuals with sleeplessness or who doze off during daytime may face a slightly increased risk of diabetes.

Previously, various studies reported on major sleep-related traits and their causal relations with type 2 diabetes, providing valuable insights into the genetic underpinnings of these associations. A recent study used a 20-year cohort analysis to determine the impact of cumulative insomnia exposure on the incidence of type 2 diabetes [26]. The study discovered a strong link between insomnia and an increased risk of acquiring type 2 diabetes, underscoring insomnia’s potential impact on metabolic health. In addition, studies also used MR to investigate the causal link between sleeplessness and the risk of developing type 2 diabetes [27]. Frequent insomnia symptoms are causally linked to higher HbA1c levels, indicating a direct relationship between insomnia and glycemic control, crucial in diabetes management [28].

Daytime dozing, or napping, has been shown to have significant implications for diabetes risk and management. Research indicates that habitual napping is associated with an increased risk of developing diabetes and poor glycemic control in individuals with diabetes. Previous studies specifically using non-self-reported data found that nap durations exceeding 60 minutes significantly elevate the risk of diabetes (OR 1.31, 95% CI 1.20 to 1.44) and are linked to poor glycemic control (OR 2.05, 95% CI 1.55 to 2.73) [29]. Long daytime naps are also associated with decreased insulin sensitivity, particularly in individuals with shorter nighttime sleep durations, suggesting that excessive napping may disrupt the sleep-wake rhythm and increase sympathetic nerve activity, thereby reducing insulin sensitivity [30]. The relationship between napping and diabetes risk appears to be influenced by factors such as body fat percentage and C-reactive protein levels, although the exact mechanisms remain unclear [31]. Furthermore, aligning with our results, a large-scale study involving 53,916 participants found that habitual daytime napping is positively associated with an increased risk of diabetes, with hazard ratios of 1.39 (95% CI 1.21–1.59) after adjusting for various socio-demographic and lifestyle factors [32].

Snoring and sleep duration have significant impacts on diabetes, with various studies highlighting their complex relationships. Snoring, a common symptom of sleep-disordered breathing, has also been linked to diabetes risk. Recent studies reported that habitual snoring is correlated with an elevated risk of developing type 2 diabetes, particularly in women [17]. The relationship between snoring and diabetes risk may be influenced by factors such as obesity, hypertension, and impaired glucose metabolism [33, 34]. Sleep duration also plays a crucial role, exhibiting a U-shaped relationship with diabetes. However, some studies indicate that the association between sleep duration and poor glycemic control is not always straightforward, as other factors like napping and overall sleep quality also contribute to glycemic outcomes [35].

It is interesting to note that the biological mechanisms linking insomnia with diabetes involve intricate pathways that include various physiological processes. Insomnia, characterized by disrupted sleep patterns, has been associated with alterations in stress response systems, particularly the hypothalamo-hypophyseal system and the pituitary-adrenal system [36]. Chronic insomnia can lead to dysregulation of cortisol, a stress hormone, which may contribute to visceral adiposity and insulin resistance, both of which are metabolic precursors to diabetes [36]. This dysregulation of cortisol may provide a biological link between insomnia and the development of type 2 diabetes. Also, daytime napping has been linked to diabetes through several biological mechanisms, as evidenced by multiple studies. One significant finding is the association between habitual daytime napping and an increased risk of type 2 diabetes (T2D), with a hazard ratio of 1.39 for those who nap regularly compared to those who do not, suggesting that napping may contribute to the development of diabetes through mechanisms related to metabolic dysregulation [32].

Genetic studies have identified 123 loci associated with daytime napping, including genes involved in arousal and sleep regulation (*HCRTR1, HCRTR2, TRPC6, PNOC*) and those suggesting an obesity-hypersomnolence pathway (*PNOC, PATJ*). These genetic links indicate that napping behavior may be influenced by genetic factors that also affect metabolic health, potentially leading to higher blood pressure and waist circumference, both of which are risk factors for diabetes [37]. On the other hand, snoring, a common symptom of sleep-disordered breathing, has been linked to diabetes risk through various mechanisms. Studies have suggested that habitual snoring is associated with increased localized carotid endothelial dysfunction due to upper airway resistance and vibrations in the pharyngeal wall, potentially contributing to cardiovascular risks, which are often comorbid with diabetes [38]. Snoring has been linked to obesity, a known risk factor for type 2 diabetes, through mechanisms that may involve increased adiposity and insulin resistance. Furthermore, both insomnia and snoring have been associated with inflammation, a common pathway implicated in the development of diabetes and its complications [39]. Additionally, oxidative stress, sympathetic nervous system activity, inflammation, adipokine levels, and insulin resistance have been suggested as potential mechanisms that connect sleep-disordered breathing events, such as snoring, to adverse health outcomes, including diabetes. [40].

This study has important strengths. This is the first exploratory study to establish the causality between a broad range of sleep-related traits and self-reported data for diabetes. Our research revealed that exposure like chronotype, insomnia, snoring, daytime dozing and diabetes outcome (self-reported) in MR studies could provide a more holistic understanding of the complex interplay between sleep disturbances and metabolic health, ultimately contributing to targeted interventions for improving diabetes outcomes. Additionally, self-reported data on sleep patterns and diabetes outcomes can capture nuanced aspects of sleep disturbances that may not be captured through objective measures alone, enhancing the understanding of the impact of insomnia on diabetes risk. Also, the study included large sample sizes and SNPs from GWASs, thereby providing sufficient statistical power for the causal association estimates.

Nevertheless, our study has several limitations. First, while acknowledging the limitations of self-reported health outcome data, its integration in MR analyses can inevitably lead to subjective bias. Secondly, despite efforts to mitigate heterogeneity, evident heterogeneity was still identified, which requires the interpretations of the findings cautiously. Thirdly, we investigate the casual link between sleep-traits and self-reported diabetes data in the European population GWAS summary data, therefore, applying this MR finding to other ethnic populations like Asian, Arab, and African people should be very cautious and additional research is required for extending these studies on different ancestries. Fourth, we used a relaxed p value threshold to confirm prior findings, therefore, may lead to false negative results. For a broader exploration of genetic associations, any significant findings should be interpreted as suggestive and warrant further validation in larger, and more robust studies. Lastly, the lack of sex stratification in the sleep-related summary data utilized for MR analysis may obscure any sex-specific relationships.

In conclusion, the study suggests a causal link between specific sleep traits, like insomnia, daytime dozing, snoring and sleep duration in individuals with diabetes by using self-reported health outcome data. Despite data limitations, such as reliance on self-reports and lack of sex stratification, the study points towards addressing these sleep disturbances may improve glycemic control and reduce other risks in diabetic populations. Moreover, this study highlights the need for these modifiable sleep traits to be added as prevention strategies for diabetes to improve public health.

## Supporting information

STROBE-MR checklist

## Abbreviations

GWAS: Genome-Wide Association Studies
MR: Mendelian randomization
SNPs: Single Nucleotide Polymorphisms
IVs: Instrumental Variables
LD: Linkage Disequilibrium
IVW: Inverse Variance Weighting
IVW-MRE: Inverse Variance Weighting -multiplicative random effects
MR-PRESSO: MR-Pleiotropy Adjusted Profile Score

## Author contributions

NAN analyzed and designed the study and wrote the initial manuscript; DV participated in the study design and critically reviewed and revised the manuscript; PVJ supervised and conceptualized the idea, attested the accuracy of the data analysis and the integrity of the data analysis. All authors reviewed and revised the manuscript.

## Acknowledgments

We greatly acknowledge UKBB for the data access and large-scale data resource collection at the MRC Integrative Epidemiology Unit (IEU) at the University of Bristol, including the collection of complete GWAS summary datasets made available as open-source files for download or by querying a database of the complete data.

## Data availability statements

The original contributions presented in the study are publicly available. The data can be found at GWAS catalog (https://gwas.mrcieu.ac.uk/).

## Supplementary material

### Supplementary file 1

Table 1: GWAS cohorts used in the study and its Descriptive information

Table 2: Sensitivity analysis before outlier correction– Pleiotropy, Heterogeneity and outlier test for Sleep-related traits with Diabetes

Table 3: Sensitivity analysis after outlier correction– Pleiotropy, Heterogeneity and outlier test for Sleep-related traits with Diabetes

Table 4: MR Analysis for Causal Associations of Sleep Traits with the Self-reported Diabetes

Table 5: Directionality analysis of sleep traits with Self-reported Diabetes

Figure S1: A. Comparison of results using different MR methods using scatter plot between Sleep duration and Self –reported Diabetes B. Forest plot of single SNP MR C. Leave-one-out sensitivity analysis D. Funnel plot

Figure S2: A. Comparison of results using different MR methods using scatter plot between Snoring and Self –reported Diabetes B. Forest plot of single SNP MR C. Leave-one-out sensitivity analysis D. Funnel plot

Figure S3: A. Comparison of results using different MR methods using scatter plot between Chronotype and Self –reported Diabetes B. Forest plot of single SNP MR C. Leave-one-out sensitivity analysis D. Funnel plot

### Supplementary file 2

All valid genetic instruments in MR analysis after QC and outlier correction for various sleep traits as exposure

## References

1. Saeedi, P., Petersohn, I., Salpea, P., Malanda, B., Karuranga, S., Unwin, N., Colagiuri, S., Guariguata, L., Motala, A.A., Ogurtsova, K., Shaw, J.E., Bright, D., Williams, R.: Global and regional diabetes prevalence estimates for 2019 and projections for 2030 and 2045: Results from the International Diabetes Federation Diabetes Atlas, 9th edition. Diabetes Res Clin Pract. 157, (2019). 10.1016/j.diabres.2019.107843

2. Mokdad, A.H., Ford, E.S., Bowman, B.A., Dietz, W.H., Vinicor, F., Bales, V.S., Marks, J.S.: Prevalence of Obesity, Diabetes, and Obesity-Related Health Risk Factors, 2001.

3. Rathmann, W., Giani, G.: Global Prevalence of Diabetes: Estimates for the Year 2000 and Projections for 2030. Diabetes Care. 27, 2568–2569 (2004). 10.2337/diacare.27.10.2568

4. Jones, S.E., Tyrrell, J., Wood, A.R., Beaumont, R.N., Ruth, K.S., Tuke, M.A., Yaghootkar, H., Hu, Y., Teder-Laving, M., Hayward, C., Roenneberg, T., Wilson, J.F., Del Greco, F., Hicks, A.A., Shin, C., Yun, C.H., Lee, S.K., Metspalu, A., Byrne, E.M., Gehrman, P.R., Tiemeier, H., Allebrandt, K. V., Freathy, R.M., Murray, A., Hinds, D.A., Frayling, T.M., Weedon, M.N.: Genome-Wide Association Analyses in 128,266 Individuals Identifies New Morningness and Sleep Duration Loci. PLoS Genet. 12, (2016). 10.1371/journal.pgen.1006125

5. Jones, S.E., van Hees, V.T., Mazzotti, D.R., Marques-Vidal, P., Sabia, S., van der Spek, A., Dashti, H.S., Engmann, J., Kocevska, D., Tyrrell, J., Beaumont, R.N., Hillsdon, M., Ruth, K.S., Tuke, M.A., Yaghootkar, H., Sharp, S.A., Ji, Y., Harrison, J.W., Freathy, R.M., Murray, A., Luik, A.I., Amin, N., Lane, J.M., Saxena, R., Rutter, M.K., Tiemeier, H., Kutalik, Z., Kumari, M., Frayling, T.M., Weedon, M.N., Gehrman, P.R., Wood, A.R.: Genetic studies of accelerometer-based sleep measures yield new insights into human sleep behaviour. Nat Commun. 10, (2019). 10.1038/s41467-019-09576-1

6. Yuan, S., Larsson, S.C.: An atlas on risk factors for type 2 diabetes: a wide-angled Mendelian randomisation study. Diabetologia. 63, 2359–2371 (2020). 10.1007/s00125-020-05253-x

7. Burgess, S., Butterworth, A., Thompson, S.G.: Mendelian randomization analysis with multiple genetic variants using summarized data. Genet Epidemiol. 37, 658– 665 (2013). 10.1002/gepi.21758

8. Mcewen, M.M., Pasvogel, A., Murdaugh, C.L.: Family Self-Efficacy for Diabetes Management: Psychometric Testing. J Nurs Meas. 32E–43E. 10.1891/1061-3749.24.1.32

9. Goldman, N., Lin, I.F., Weinstein, M., Lin, Y.H.: Evaluating the quality of self-reports of hypertension and diabetes. J Clin Epidemiol. 56, 148–154 (2003). 10.1016/S0895-4356(02)00580-2

10. Saddki, N., Hashim, M.I.B.M., Mohamad, N.: Factors Associated with Oral Health Literacy among Patients with Type 2 Diabetes Mellitus Attending Hospital Universiti Sains Malaysia. Pesqui Bras Odontopediatria Clin Integr. 22, (2022). 10.1590/pboci.2022.056

11. Schneider, A.L.C., Pankow, J.S., Heiss, G., Selvin, E.: Validity and reliability of self-reported diabetes in the atherosclerosis risk in communities study. Am J Epidemiol. 176, 738–743 (2012). 10.1093/aje/kws156

12. Hemani, G., Zheng, J., Elsworth, B., Wade, K.H., Haberland, V., Baird, D., Laurin, C., Burgess, S., Bowden, J., Langdon, R., Tan, V.Y., Yarmolinsky, J., Shihab, H.A., Timpson, N.J., Evans, D.M., Relton, C., Martin, R.M., Davey Smith, G., Gaunt, T.R., Haycock, P.C.: The MR-Base platform supports systematic causal inference across the human phenome. (2018). 10.7554/eLife.34408.001

13. Skrivankova, V.W., Richmond, R.C., Woolf, B.A.R., Yarmolinsky, J., Davies, N.M., Swanson, S.A., VanderWeele, T.J., Higgins, J.P.T., Timpson, N.J., Dimou, N., Langenberg, C., Golub, R.M., Loder, E.W., Gallo, V., Tybjaerg-Hansen, A., Davey Smith, G., Egger, M., Richards, J.B.: Strengthening the Reporting of Observational Studies in Epidemiology Using Mendelian Randomization: The STROBE-MR Statement. JAMA. 326, 1614–1621 (2021). 10.1001/jama.2021.18236

14. Ai, Q., Yang, B.: Are inflammatory bowel diseases associated with an increased risk of COVID-19 susceptibility and severity? A two-sample Mendelian randomization study. Front Genet. 14, (2023). 10.3389/fgene.2023.1095050

15. Bowden, J., Smith, G.D., Burgess, S.: Mendelian randomization with invalid instruments: Effect estimation and bias detection through Egger regression. Int J Epidemiol. 44, 512–525 (2015). 10.1093/ije/dyv080

16. Wan, A., Zhao, W.D., Tao, J.H.: Causal effects of systemic lupus erythematosus on endometrial cancer: A univariable and multivariable Mendelian randomization study. Front Oncol. 12, (2022). 10.3389/fonc.2022.930243

17. Yuan, Y., Zhang, F., Qiu, J., Chen, L., Xiao, M., Tang, W., Luo, Q., Ding, X., Tang, X.: Association Between Snoring and Diabetes Among Pre-and Postmenopausal Women. Int J Gen Med. 15, 2491–2499 (2022). 10.2147/IJGM.S352593

18. Yang, Y., Fan, J., Shi, X., Wang, Y., Yang, C., Lian, J., Wang, N., Zhao, C., Zhao, Y., Jia, X.: Causal associations between sleep traits and four cardiac diseases: a Mendelian randomization study. ESC Heart Fail. 9, 3160–3166 (2022). 10.1002/ehf2.14016

19. Ma, K., Song, P., Liu, Z., Yang, L., Wang, L., Zhou, J., Chen, J., Dong, Q.: Genetic evidence suggests that depression increases the risk of erectile dysfunction: A Mendelian randomization study. Front Genet. 13, (2022). 10.3389/fgene.2022.1026227

20. Jiang, H., Li, Y., Shen, J., Lin, H., Fan, S., Qiu, R., He, J., Lin, E., Chen, L.: Cigarette smoking and thyroid cancer risk: A Mendelian randomization study. Cancer Med. 12, 19866–19873 (2023). 10.1002/cam4.6570

21. Luo, P., Hou, W., Xu, K., Liu, L., Xu, P.: The Causal Relationship Between Rheumatoid Arthritis and Pneumonia: A Mendelian Randomization Study. (2022). 10.21203/rs.3.rs-2022175/v1

22. Wan, X., Xie, J., Yang, M., Yu, H., Hou, W., Xu, K., Wang, J., Xu, P.: Does Having Rheumatoid Arthritis Increase the Dose of Depression Medications? A Mendelian Randomization Study. J Clin Med. 12, (2023). 10.3390/jcm12041405

23. Karasik, D., Jeong, S., Jie Xie, W., Shen, H., Chen, S., M-g, D.: The causal impact of childhood obesity on bone mineral density and fracture in adulthood: A two-sample Mendelian randomization study.

24. Hemani, G., Bowden, J., Davey Smith, G.: Evaluating the potential role of pleiotropy in Mendelian randomization studies, (2018)

25. Mazidi, M., Dehghan, A., Banach, M.: Genetically higher level of mannose has no impact on cardiometabolic risk factors: Insight from mendelian randomization. Nutrients. 13, (2021). 10.3390/nu13082563

26. Green, M.J., Espie, C.A., Popham, F., Robertson, T., Benzeval, M.: Insomnia symptoms as a cause of type 2 diabetes Incidence: A 20 year cohort study. BMC Psychiatry. 17, (2017). 10.1186/s12888-017-1268-4

27. Gao, X., Sun, H., Zhang, Y., Liu, L., Wang, J., Wang, T.: Investigating Causal Relations Between Sleep-Related Traits and Risk of Type 2 Diabetes Mellitus: A Mendelian Randomization Study. Front Genet. 11, (2020). 10.3389/fgene.2020.607865

28. Tian, M., Ma, H., Shen, J., Hu, T., Cui, H., Huangfu, N.: Causal association between sleep traits and the risk of coronary artery disease in patients with diabetes. Front Cardiovasc Med. 10, (2023). 10.3389/fcvm.2023.1132281

29. Liu, M., Wang, S., Sun, Y., Zhou, F., Sun, H.: Relationship between daytime napping with the occurrence and development of diabetes: a systematic review and meta-analysis. BMJ Open. 13, (2023). 10.1136/bmjopen-2022-068554

30. Kakutani-Hatayama, M., Kadoya, M., Morimoto, A., Miyoshi, A., Kosaka-Hamamoto, K., Kanzaki, A., Konishi, K., Kusunoki, Y., Syoji, T., Koyama, H.: Excessive daytime napping independently associated with decreased insulin sensitivity in cross-sectional study – Hyogo Sleep Cardio-Autonomic Atherosclerosis cohort study. Front Endocrinol (Lausanne). 14, (2023). 10.3389/fendo.2023.1211705

31. Zhou, R., Chen, H., Huang, Y.-N., Zhong, Q., Huang, R., Liu, H., Zheng, J., Wu, X.-B.: The association between daytime napping and risk of type 2 diabetes is modulated by inflammation and adiposity: Evidence from 435□342 UK□Biobank participants. J Diabetes. 15, 496–507 (2023). 10.1111/1753-0407.13387

32. Wang, H., Chen, L., Shen, D., Cao, Y., Zhang, X., Xie, K., Wang, C., Zhu, S., Guo, Y., Fiona, B., Yu, M., Chen, Z., Li, L.: Association of daytime napping in relation to risk of diabetes: evidence from a prospective study in Zhejiang, China. Nutr Metab (Lond). 18, 1–8 (2021). 10.1186/S12986-021-00545-4

33. Wei, Y., Zheng, B., Fan, J., Lv, J., Guo, Y., Bian, Z., Du, H., Yang, L., Chen, Y., Chen, J., Zhong, X., Chen, J., Chen, Z., Yu, C., Li, L.: Habitual snoring, adiposity measures and risk of type 2 diabetes in 0.5 million Chinese adults: A 10-year cohort. BMJ Open Diabetes Res Care. 8, (2020). 10.1136/bmjdrc-2019-001015

34. Ramirez, J., Nguyen Thi Phuong, M., Nguyen Ngoc Phuong, T., Yue, W., Copyright, fnagi, Zhang, Y., Zhang, T., Xia, X., Hu, Y., Zhang, C., Liu, R., Yang, Y., Li, X.: OPEN ACCESS EDITED BY The relationship between sleep quality, snoring symptoms, night shift and risk of stroke in Chinese over rr years old.

35. Bawadi, H., Sada, A. Al, Mansoori, N. Al, Mannai, S. Al, Hamdan, A., Shi, Z., Kerkadi, A.: Sleeping Duration, Napping and Snoring in Association with Diabetes Control among Patients with Diabetes in Qatar. Int J Environ Res Public Health. 18, 4017 (2021). 10.3390/IJERPH18084017

36. Joseph, J.J., Golden, S.H.: Cortisol dysregulation: the bidirectional link between stress, depression, and type 2 diabetes mellitus, (2017)

37. Dashti, H.S., Daghlas, I., Daghlas, I., Lane, J.M., Lane, J.M., Huang, Y., Udler, M.S., Wang, H., Ollila, H., Jones, S.E., Kim, J., Wood, A.R., Weedon, M.N., Aslibekyan, S., Garaulet, M., Saxena, R., Saxena, R.: Genetic determinants of daytime napping and effects on cardiometabolic health. Nat Commun. 12, 1–15 (2021). 10.1038/S41467-020-20585-3

38. Sands, M., Loucks, E.B., Lu, B., Carskadon, M.A., Sharkey, K., Stefanick, M., Ockene, J., Shah, N., Hairston, K.G., Robinson, J., Limacher, M., Hale, L., Eaton, C.B.: Self-reported snoring and risk of cardiovascular disease among postmenopausal women (from the Women’s Health Initiative). American Journal of Cardiology. 111, 540–546 (2013). 10.1016/j.amjcard.2012.10.039

39. Hackett, R.A., Steptoe, A.: Psychosocial Factors in Diabetes and Cardiovascular Risk, (2016)

40. Izci-Balserak, B., Pien, G.W.: Sleep-disordered breathing and pregnancy: Potential mechanisms and evidence for maternal and fetal morbidity, (2010)

