## Supplementary File 1 for "Investigating the Causal Relationship Between Sleep-Related Traits and Self-Reported Diabetes: A Mendelian Randomization Study"

**Investigating the Causal Relationship Between Sleep-Related Traits and Self-Reported Diabetes: A Mendelian Randomization Study**

**Table 1** GWAS cohorts used in the study and its Descriptive information

**Table 2** Sensitivity analysis before outlier correction– Pleiotropy, Heterogeneity and outlier test for Sleep-related traits with Diabetes

**Table 3** Sensitivity analysis after outlier correction– Pleiotropy, Heterogeneity and outlier test for Sleep-related traits with Diabetes

**Table 4**: MR Analysis for Causal Associations of Sleep Traits with the Self-reported Diabetes

**Table 5** Directionality analysis of sleep traits with Self-reported Diabetes

**Figure S1: A.** Comparison of results using different MR methods using scatter plot between Sleep duration and Self –reported Diabetes B. Forest plot of single SNP MR **C.** Leave-one-out sensitivity analysis D. Funnel plot

**Figure S2: A.** Comparison of results using different MR methods using scatter plot between Snoring and Self –reported Diabetes B. Forest plot of single SNP MR **C.** Leave-one-out sensitivity analysis D. Funnel plot

**Figure S3: A.** Comparison of results using different MR methods using scatter plot between Chronotype and Self –reported Diabetes B. Forest plot of single SNP MR **C.** Leave-one-out sensitivity analysis D. Funnel plot

**Table 1** GWAS cohorts used in the study and its Descriptive information

| **Phenotype** | **GWAS ID** | **Web Source** | **Sample Size** | **Number of SNPs** | **Consortium** | **Population** |
| --- | --- | --- | --- | --- | --- | --- |
| Sleep duration | ukb-a-9 | <https://gwas.mrcieu.ac.uk/datasets/ukb-a-9/> | 335,410 | 10,894,596 | Neale Lab | European |
| Insomnia | ukb-b-3957 | <https://gwas.mrcieu.ac.uk/datasets/ukb-b-3957/> | 462,341 | 9,851,867 | MRC-IEU | European |
| Chronotype | ukb-b-4956 | <https://gwas.mrcieu.ac.uk/datasets/ukb-b-4956/> | 413,343 | 9,851,867 | MRC-IEU | European |
| Snoring | ukb-b-17400 | <https://gwas.mrcieu.ac.uk/datasets/ukb-b-17400/> | 430,438 | 9,851,867 | MRC-IEU | European |
| Daytime  dozing | ukb-b-5776 | [Trait: Daytime dozing / sleeping (narcolepsy) - IEU OpenGWAS project (mrcieu.ac.uk)](https://gwas.mrcieu.ac.uk/datasets/ukb-b-5776/) | 460,913 | 9,851,867 | MRC-IEU | European |
| Self-reported  Diabetus^1^ | ukb-b-12948 | <https://gwas.mrcieu.ac.uk/datasets/ukb-b-12948/> | 462,933 | 9,851,867 | MRC-IEU | European |

^1^Non-cancer illness code: Self-report, UK Biobank data

**Table 2** Sensitivity analysis before outlier correction– Pleiotropy, Heterogeneity and outlier test for Sleep-related traits with Diabetes

| **Exposure** | **Pleiotropy Test**  **(MR-Egger)** | | **Heterogeneity Test**  **(IVW)** | | **Outlier Test**  **(MR-PRESSO)** |
| --- | --- | --- | --- | --- | --- |
|  | **Egger Intercept** | ***p-value*** | **Q** | **P-value** | **Outliers** |
| Sleep duration | -0.0008 | 0.114 | 140 | 1.106×10^-12^ | rs3751813, rs448231,  rs62158206, rs925872 |
| Insomnia | 0.0003 | 0.422 | 100 | 2.023×10^-7^ | rs2014830 |
| Chronotype | -0.00003 | 0.888 | 420 | 5.225×10^−27^ | rs12140153, rs1421085,  rs2291589, rs4237555,  rs4241964, rs4729854 |
| Snoring | 0.00005 | 0.932 | 83 | 1.017×10^−4^ | rs1558901, rs2307111 |
| Daytime dozing | -0.0012 | 0.101 | 122 | 4.846×10^−13^ | rs12140153, rs4665972,  rs55767040 |

**Table 3** Sensitivity analysis after outlier correction– Pleiotropy, Heterogeneity and outlier test for Sleep-related traits with Diabetes

| **Exposure** | **Pleiotropy Test**  **(MR-Egger)** | | **Heterogeneity Test**  **(IVW)** | | **Outlier Test**  **(MR-PRESSO)** |
| --- | --- | --- | --- | --- | --- |
|  | **Egger Intercept** | ***p-value*** | **Q** | **P-value** | **Outliers** |
| Sleep duration | 0.000262 | 0.275 | 58 | 2.3×10^−2^ | No significant outliers |
| Insomnia | 0.000293 | 0.396 | 96 | 1×10^−4^ | No significant outliers |
| Chronotype | -0.000117 | 0.484 | 254 | 1×10^−7^ | No significant outliers |
| Snoring | 0.000007 | 0.989 | 66 | 5×10^−3^ | No significant outliers |
| Daytime dozing | -0.000004 | 0.995 | 43 | 2.3×10^−2^ | No significant outliers |

**Table 4**: MR Analysis for Causal Associations of Sleep Traits with the Self-reported Diabetes

| **Nos** | **Exposure** | **N SNP** | **Method** | **Beta** | **Pval** | **Odd Ratio**  **(95% CI)** |
| --- | --- | --- | --- | --- | --- | --- |
| 1 | Sleep Duration | 40 | Inverse variance weighted | -0.004 | 0.24982 | 0.996(0.990, 1.003) |
|  |  |  | Inverse variance weighted   (multiplicative random effects) | -0.004 | 0.24982 | 0.996(0.990, 1.003) |
|  |  |  | Maximum likelihood | -0.004 | 0.17033 | 0.996(0.991, 1.002) |
|  |  |  | MR Egger | -0.024 | 0.20300 | 0.976(0.941, 1.013) |
|  |  |  | Weighted median | -0.008 | 0.05335 | 0.992(0.984, 1.000) |
| 2 | Insomnia | 38 | Inverse variance weighted | 0.020 | 0.05490 | 1.020(0.999, 1.042) |
|  |  |  | Inverse variance weighted   (multiplicative random effects) | 0.020 | 0.05490 | 1.020(0.999, 1.042) |
|  |  |  | Maximum likelihood | 0.021 | 0.00186 | 1.021(1.008, 1.035) |
|  |  |  | MR Egger | -0.009 | 0.79839 | 0.991(0.929, 1.058) |
|  |  |  | Weighted median | 0.011 | 0.33372 | 1.011(0.989, 1.033) |
| 3 | Chronotype | 148 | Inverse variance weighted | 0.001 | 0.65465 | 1.001(0.995, 1.008) |
|  |  |  | Inverse variance weighted  (multiplicative random effects) | 0.001 | 0.65465 | 1.001(0.995, 1.008) |
|  |  |  | Maximum likelihood | 0.001 | 0.55410 | 1.001(0.997, 1.006) |
|  |  |  | MR Egger | 0.008 | 0.42302 | 1.008(0.988, 1.029) |
|  |  |  | Weighted median | -0.001 | 0.87111 | 0.999(0.992, 1.007) |
| 4 | Snoring | 41 | Inverse variance weighted | -0.048 | 0.00013 | 1.007(0.998, 1.016) |
|  |  |  | Inverse variance weighted   (multiplicative random effects) | -0.048 | 0.00013 | 1.007(0.998, 1.016) |
|  |  |  | Maximum likelihood | -0.048 | 0.000002 | 1.008(0.998, 1.017) |
|  |  |  | MR Egger | -0.049 | 0.47445 | 1.064(1.013, 1.117) |
|  |  |  | Weighted median | -0.034 | 0.03162 | 1.003(0.991, 1.017) |
| 5 | Daytime dozing | 28 | Inverse variance weighted | 0.028 | 0.05351 | 1.028(0.999, 1.057) |
|  |  |  | Inverse variance weighted   (multiplicative random effects) | 0.028 | 0.05351 | 1.028(0.999, 1.057) |
|  |  |  | Maximum likelihood | 0.028 | 0.01374 | 1.029(1.006, 1.052) |
|  |  |  | MR Egger | 0.028 | 0.73159 | 1.029(0.877, 1.206) |
|  |  |  | Weighted median | 0.020 | 0.25953 | 1.020(0.985, 1.057) |

**Table 5** Directionality analysis of sleep traits with Self-reported Diabetes

| **Exposure** | **snp_r2. exposure** | **snp_r2. outcome** | **correct_causal_direction** | **steiger_pval** |
| --- | --- | --- | --- | --- |
| Sleep duration | 0.0050 | 0.0003 | TRUE | 1.27E-121 |
| Insomnia | 0.0041 | 0.0002 | TRUE | 7.93E-121 |
| Chronotype | 0.0183 | 0.0006 | TRUE | 0 |
| Snoring | 0.0041 | 0.0005 | TRUE | 1.85E-88 |
| Daytime dozing | 0.0030 | 0.0003 | TRUE | 8.80E-75 |

**A. B.**

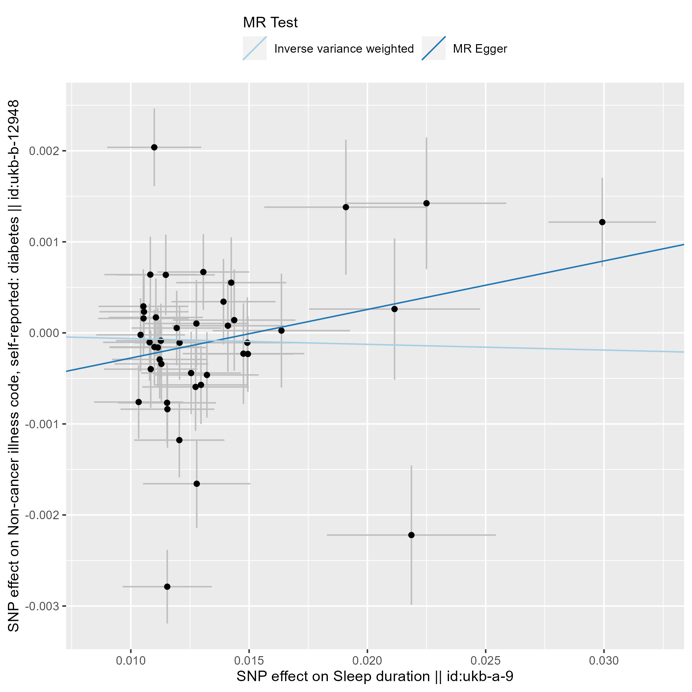

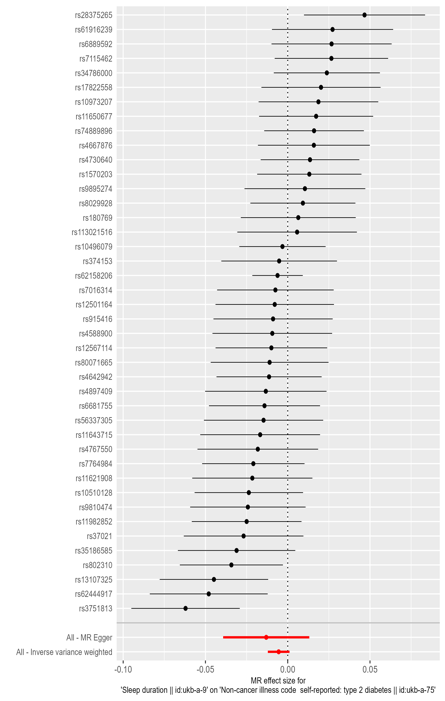

**C. D.**

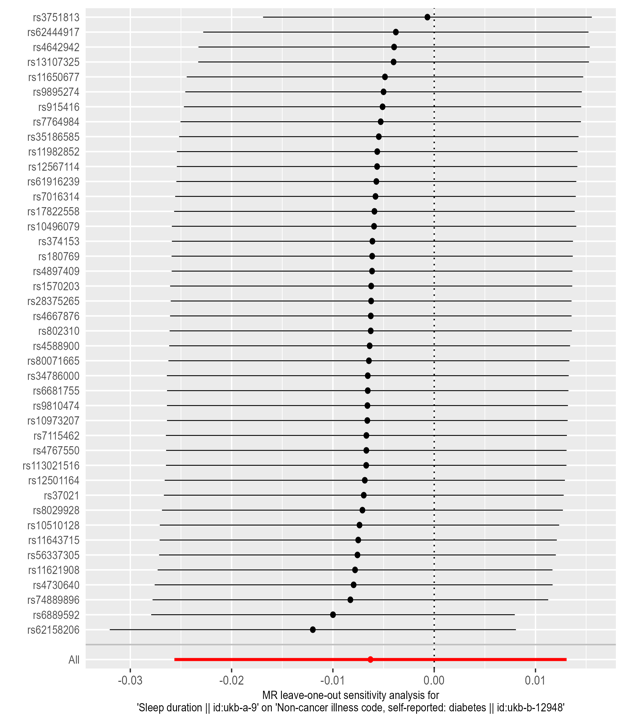

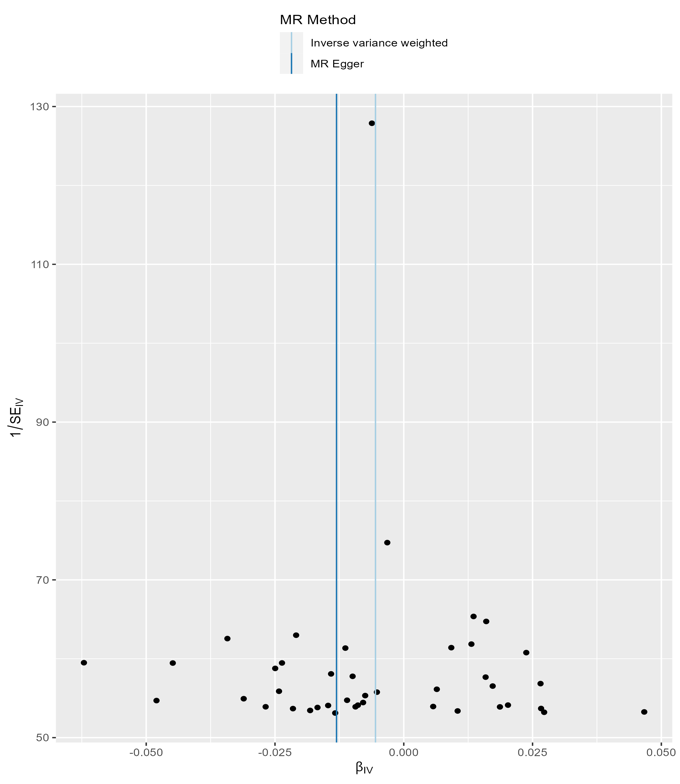
**Fig S1: A.** Comparison of results using different MR methods using scatter plot between Sleep duration and Self –reported Diabetes B. Forest plot of single SNP MR **C.** Leave-one-out sensitivity analysis D. Funnel plot

**A. B.**

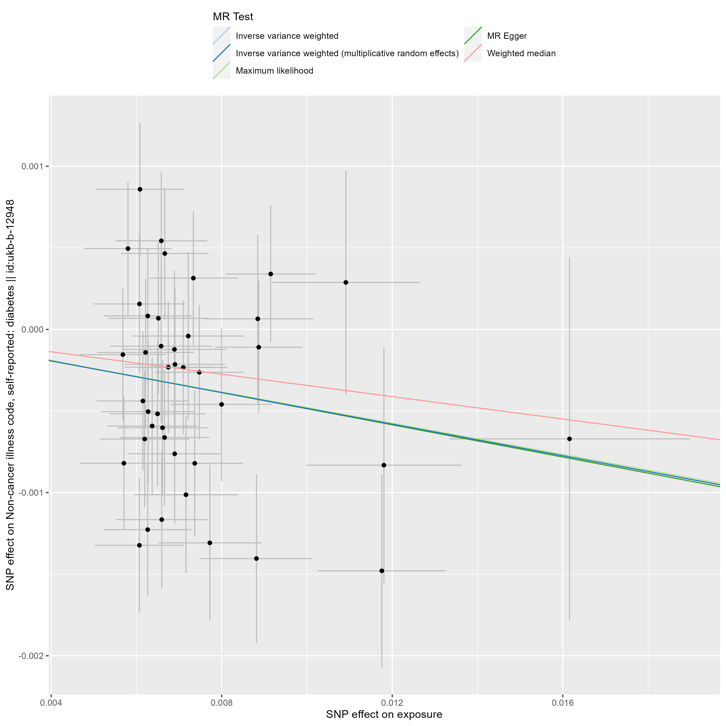

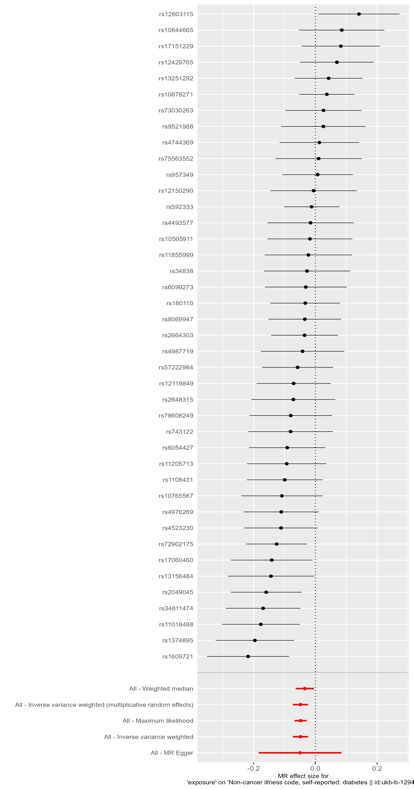

**C. D.**

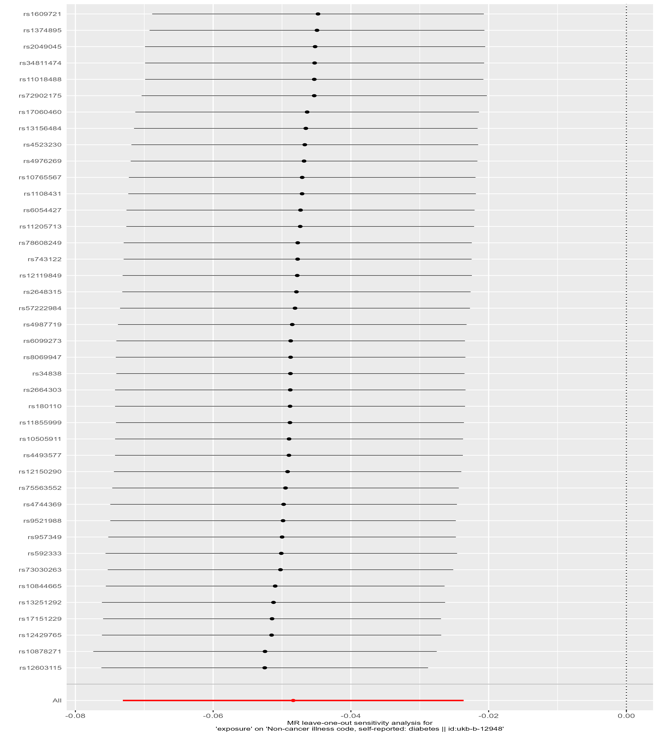

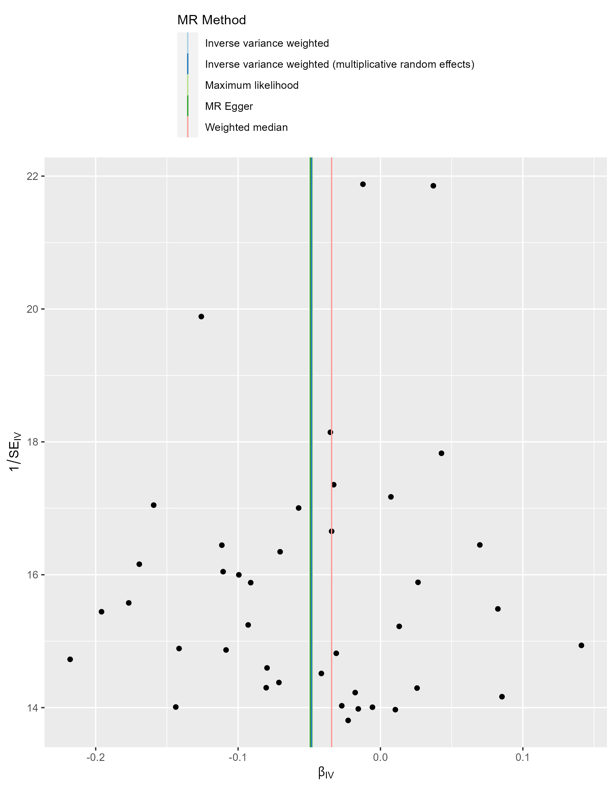

**Fig S2: A.** Comparison of results using different MR methods using scatter plot between Snoring and Self –reported Diabetes B. Forest plot of single SNP MR **C.** Leave-one-out sensitivity analysis D. Funnel plot

**A. B.**

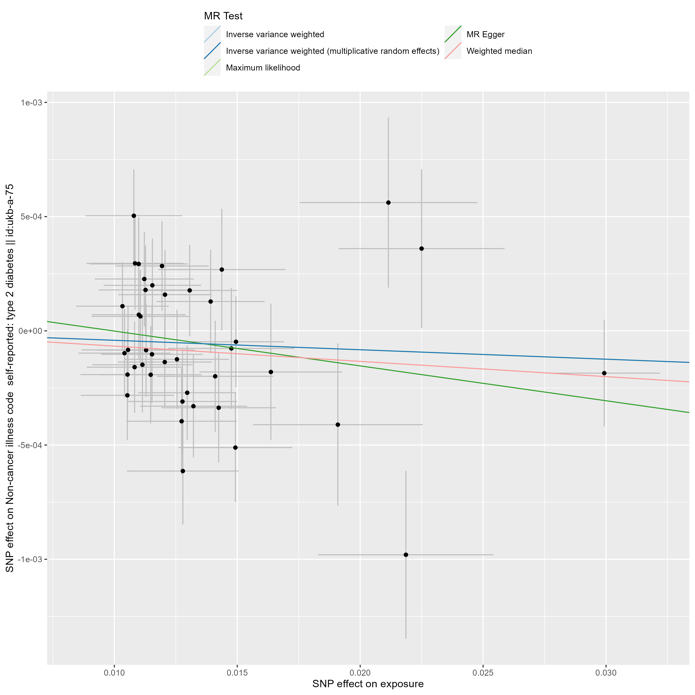

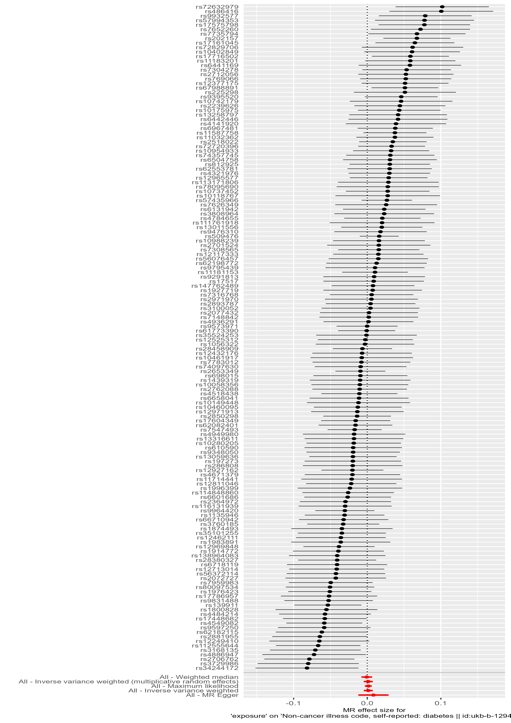

**C. D.**

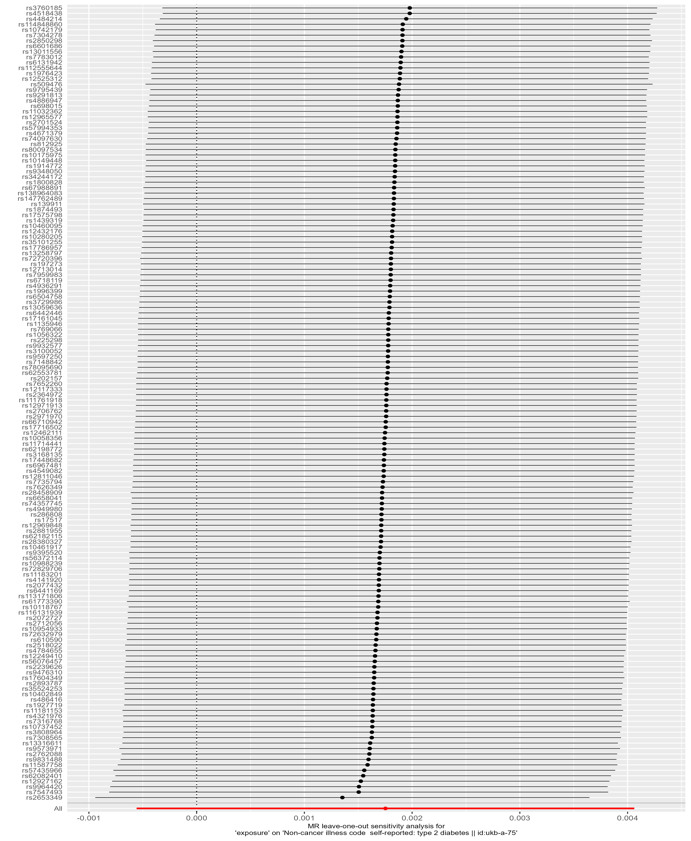

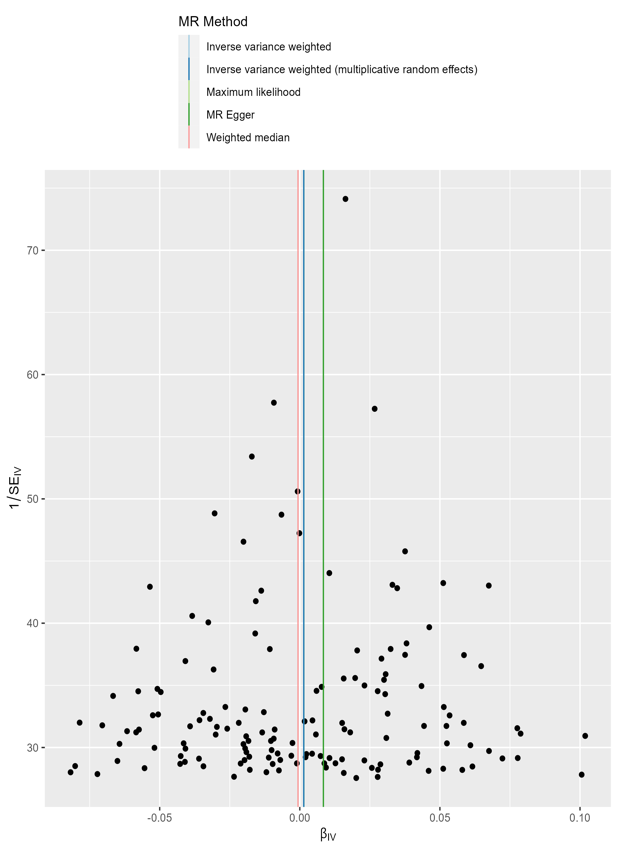
**Fig S3: A.** Comparison of results using different MR methods using scatter plot between Chronotype and Self –reported Diabetes B. Forest plot of single SNP MR **C.** Leave-one-out sensitivity analysis D. Funnel plot
